# Prevalence and effect of bacterial co-infections on clinical outcomes in hospitalized COVID-19 patients at a tertiary care centre of India

**DOI:** 10.1101/2021.08.06.21261695

**Authors:** Ashutosh Pathak, Sweta Singh, Sanjay Singh, Sangram Singh Patel, Nida Fatima, Atul Garg, Ujjala Ghoshal, Chinmoy Sahu

## Abstract

**Background:** Bacterial co-infections are a leading cause of morbidity and mortality during viral infections including COVID-19. Systematic testing of COVID-19 patients having bacterial co-infections is essential to select the correct antibiotic for treatment in order to reduce mortality and also prevent spread of antimicrobial resistance (AMR). The present study aims to evaluate the prevalence, demographic parameters, antibiotic sensitivity patterns and outcomes in hospitalized COVID-19 patients with bacterial co-infections.

**Methods:** A total of 1019 COVID-19 patients were selected for the study. We analyzed the prevalence, antibiotic sensitivity pattern and clinical outcomes in COVID-19 patients having bacterial co-infections.

**Results:** Out of a total 1019 COVID-19 patients screened, 5.2% of patients demonstrated clinical signs of bacterial co-infection. Bacteremia was found in majority of the patients followed by respiratory and urinary infections. *Escherichia coli, Pseudomonas aeruginosa* and *Klebsiella spp*. were most common isolates among the Gram-negative and Coagulase-negative Staphylococci (CONS) and *Staphylococcus aureus* among the Gram-positive bacterial infections. Antibiotic sensitivity profiling revealed that colistin, imipenem and fosfomycin were the most effective drugs against the Gram-negative isolates while vancomycin, teicoplanin and doxycycline against the Gram-positive isolates. Analysis of clinical outcomes revealed that the mortality rate was higher (39%) among the patients with bacterial co-infections as compared to the group without co-infection (17%).

**Conclusions:** This study reveals that the rate of bacterial co-infections is significantly increasing among COVID-19 patients and leading to increase in mortality. Systematic testing of bacterial co-infections is therefore essential in COVID-19 patients for better clinical outcomes and to reduce AMR.

## Introduction

The human race has witnessed the emergence of four severe viral outbreaks in the last two decades: the Severe Acute Respiratory Syndrome Coronavirus (SARS-CoV) epidemic in 2002, the influenza A H1N1 pandemic in 2012, Middle East Respiratory Syndrome (MERS) outbreak in 2009 and most recently the COVID-19 pandemic [1]. COVID-19 has emerged as a global pandemic affecting millions of people world-wide and is proving to be a greater danger than MERS and SARS Coronaviruses [2]. A plethora of symptoms have been described in the past few months, clearly indicating that COVID-19 is a complex disease, which in no way consists only of a respiratory infection. It presents with a variety of unspecific symptoms so that the differential diagnosis encompasses a wide range of infections.Most common symptoms of COVID-19 include respiratory symptoms like cough, sputum, shortness of breath, fever; musculoskeletal symptoms like myalgia, joint pain, headache, fatigue; enteric symptoms like abdominal pain, vomiting, diarrhea and mucocutaneous symptoms (less common) [3]. Lung infections may progress in a few cases to ARDS (Acute respiratory distress syndrome), shock and death. Cytokine storm, immune dysregulation and various viral evasion mechanisms in the presence of various co-morbid conditions has contributed to fatal outcomes in the COVID-19 patients [4].

Super-infection with bacterial pathogens has been identified in various viral respiratory illnesses in the past and contributed to significant rates of morbidity and mortality [5–7]. Mortality has been reported to be as high as 20-30% in such cases of super-infection [7, 8]. Existence of bacterial co-pathogens in such respiratory patients leads to increased hospital stay, greater chances of acquiring nosocomial infections and overall increase in cost of hospital stay [9]. SARS-CoV-2 is a new virus with still limited knowledge about its pathogenesis and clinical manifestations. Hence, there are several areas of knowledge gap regarding this novel Coronavirus. One such area of potential research and studies is about the co-existence, prevalence and incidence of bacterial pathogens in SARS-CoV-2[10, 11]. Various anti-viral and immuno-modulatory agents are being tried in COVID-19 hospitalized patients, and several others are in experimental phase and under trials. Antibiotics are of no use in such patients but are routinely prescribed due to the potential risk of secondary bacterial infections [10]. Various studies have advocated the use of antibiotics in patients suffering from respiratory symptoms of COVID-19 [12, 13]. However, such irrational and unguided antibiotic use ultimately leads to emergence of antimicrobial resistance [11].

Hence, there is a dire need to study bacterial co-infections in COVID-19 patients and understand the exact prevalence of such co-pathogens for the proper and rational use of antibiotics. This study was planned with the aim to study the prevalence, demographic parameters, risk factors, antibiotic sensitivity patterns and outcomes in hospitalized COVID-19 patients with bacterial co-infections.

## Material and Methods

### Place & Duration of Study

The study was conducted at a tertiary care hospital of Northern India from March 2020 to August 2020 (six months) during the COVID-19 pandemic.

### Study design

During the time span of six months; a total of 1019 hospitalized COVID-19 patients were included in the study. The patients were analyzed for the presence of bacterial co-infection as determined by the presence of characteristic clinical features and culture positive blood, respiratory, urine or pus/aspiration samples. The patients who were confirmed to have bacterial co-infection were further analyzed. We collected following information on COVID-19 patients with bacterial co-infection: demographic details of the admitted patients, presence of co-morbidities (if any), date of admission and culture-positive results, day of hospitalization on which bacterial culture was sent, antibiotic susceptibility profiles of the bacterial isolates, length of hospital stay and clinical outcomes in the patients.

### Molecular testing for SARS CoV-2

Two swabs, one oropharyngeal and the other deep nasal swab were collected from the patients suspected of having SARS CoV-2and transported in viral transport media (VTM) to the laboratory. Viral RNA extraction was done using the QIAampViral RNA kit (Qiagen Inc., USA) according to the manufacturer’s instructions. Real Time PCR was performed by Applied Biosystems TaqPath COVID-19 kit (Thermo Fisher Scientific, USA) according to the manufacturer’s instructions.

### Bacterial culture & processing

All the specimens (blood, respiratory, urine or pus /aspiration) were cultured on both MacConkey and blood agar plates according to standard microbiological techniques. Further, colonies were isolated and subcultures done accordingly. The bacterial isolates were first identified using routine staining and biochemical tests [14]. The identities of bacteria were further confirmed by MALDI-TOF MS and Vitek 2 system (Biomerieux Inc., France), an automated identification and susceptibility testing system[15].

### Antibiotic susceptibility testing

Antibiotic susceptibility testing was done by Kirby-Bauer’s disk diffusion method on Muller Hinton agar [16]. Results were interpreted based on Clinical and Laboratory Standards Institute (CLSI) guidelines [17].

### Patient follow-up

Organism profiles, sensitivity and other study parameters were kept in computer database along with particulars of the patients. WHO classified the drug usage in hospitalized patients as per the AWaRe criteria : Access, Watch and Reserve. This criteria was used to study the overall trend of antibiotic usage in the patients. Follow up was planned in the suspected cases for any repeat culture, outcomes & associated co-morbidities.

### Statistical Analysis

The results were analyzed using the SPSS version 22 software (SPSS Inc., Chicago, IL, USA). The frequencies are shown with 95% confidence intervals (95% CI). The Chi-square and Mann-Whitney U test was used to analyse the statistically significant variables. The statistically significant values were considered as p-value<0.05.

## Results

A total of 1019 patients tested positive for COVID-19 in a span of six months and were hospitalized at our COVID hospital. Out of these hospitalized COVID-19 patients, 5.2% (n=53/1019) showed clinical signs of bacterial co-infection. Out of the total number of patients having confirmed diagnosis of bacterial co-infection, a majority (66%; n=35/53) acquired infections in the hospital (>48 hours of admission). The mean age of the patients was 68 years (SD=57 ± 16) and it ranged from 8-75 years. Males comprised the majority of the cases (61 %; n=32/53) as compared to females. Various co-morbidities in the patients were also studied; hypertension (61%; n= 32/53) and diabetes mellitus (58%; n=31/53) emerged as the most prevalent medical conditions. COPD/Asthma (27%; n=14/53), chronic kidney disease (CKD) (23%; n=12/53) and chronic liver diseases (CLD) (19%; n=10/53) were the other major co-morbidities among the COVID-19 patients.

### Etiology of the bacterial co-infections

Among the various samples collected from COVID-19 patients suffering from co-infections, blood was received from majority of the cases (37%; n=20/53). Urine (31%; n=16/53), respiratory specimens (28%; n=14/53) and pus/aspirated fluid (4%; n=3/53) contributed for the other bacteriological samples. Bacteremia was seen in majority of the COVID-19 positive patients; followed by respiratory and urinary symptoms.

*E. coli* was the common isolate (21%; n=11/53), followed by *Pseudomonas aeruginosa* (13.2%; n=7/53) and *Klebsiella spp*. (11.3%; n=6/53) among the Gram-negative organisms. *Burkholderia cepacia, Stenotrophomonas maltophila, Morganella morganii* and *Proteus mirabilis* were isolated from one patient each. On the other hand; Coagulase-negative *Staphylococci* (CONS) (18.8 %; n=10/53) and *Staphylococcus aureus* (17%; n=9/53) formed the majority of isolates in Gram-positive organisms. Methicillin resistant *Staphylococcus aureus* (MRSA) was seen in 4, Methicillin resistant coagulase-negative *Staphylococci* (MRCONS) in 7, *Enterococcus* spp. in 5 and *Streptococcus* spp. in 1 case.

### Antibiotic sensitivity profile of bacterial isolates

Antibiotic sensitivity testing revealed colistin (99%), imipenem (78%) and fosfomycin (95%) (only in urinary isolates) as the most effective drugs against Gram-negative isolates whereas vancomycin (100%), teicoplanin (99%) and doxycycline (71%) emerged as the most potent drugs for Gram-positive bacterial infections. Detailed antibiotic sensitivity patterns for both Gram-negative and Gram-positive isolates are summarized in Fig 1 & 2 respectively.

**Fig 1:**
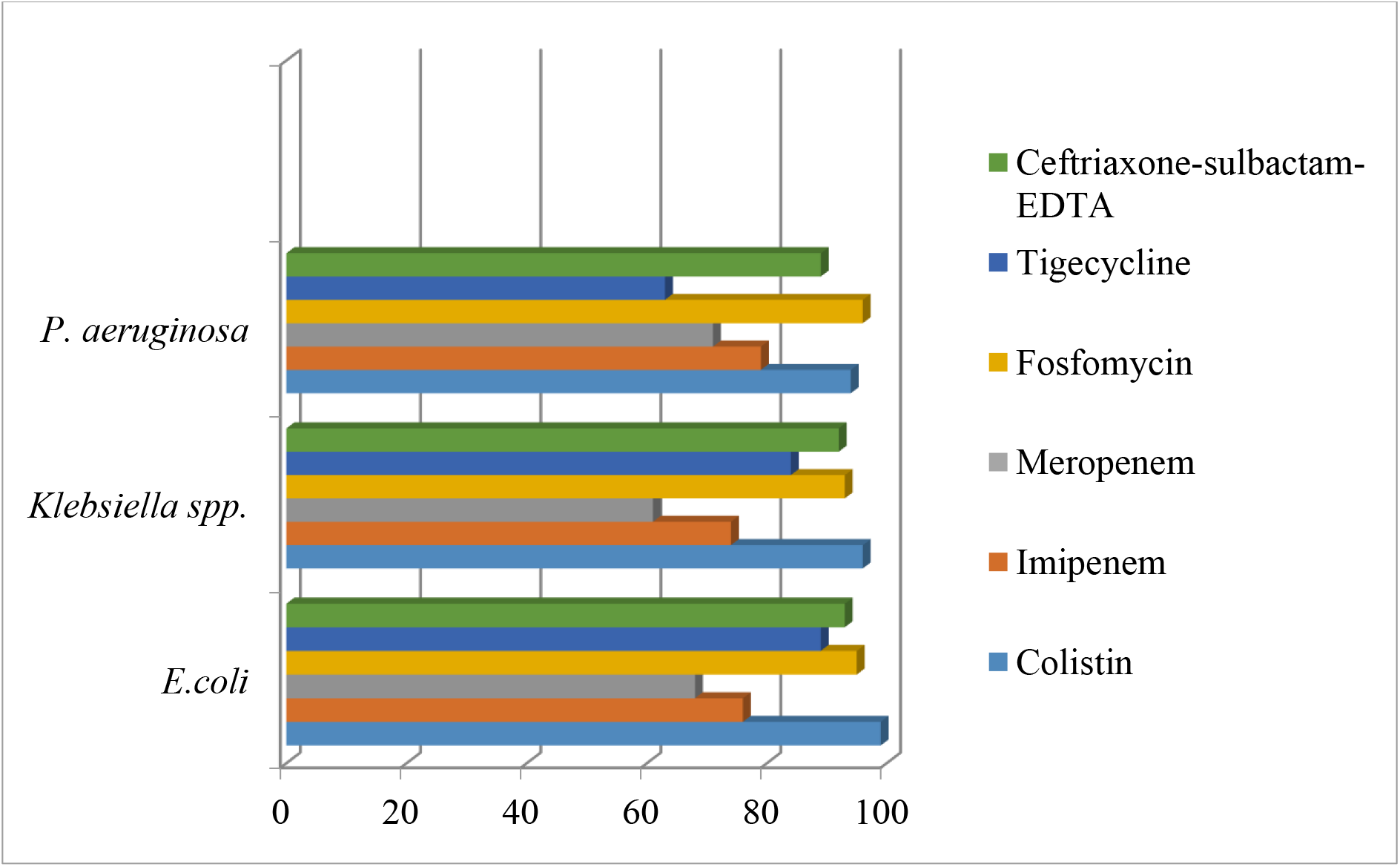
Antibiotic sensitivity pattern in Gram-negative isolates.

**Fig 2:**
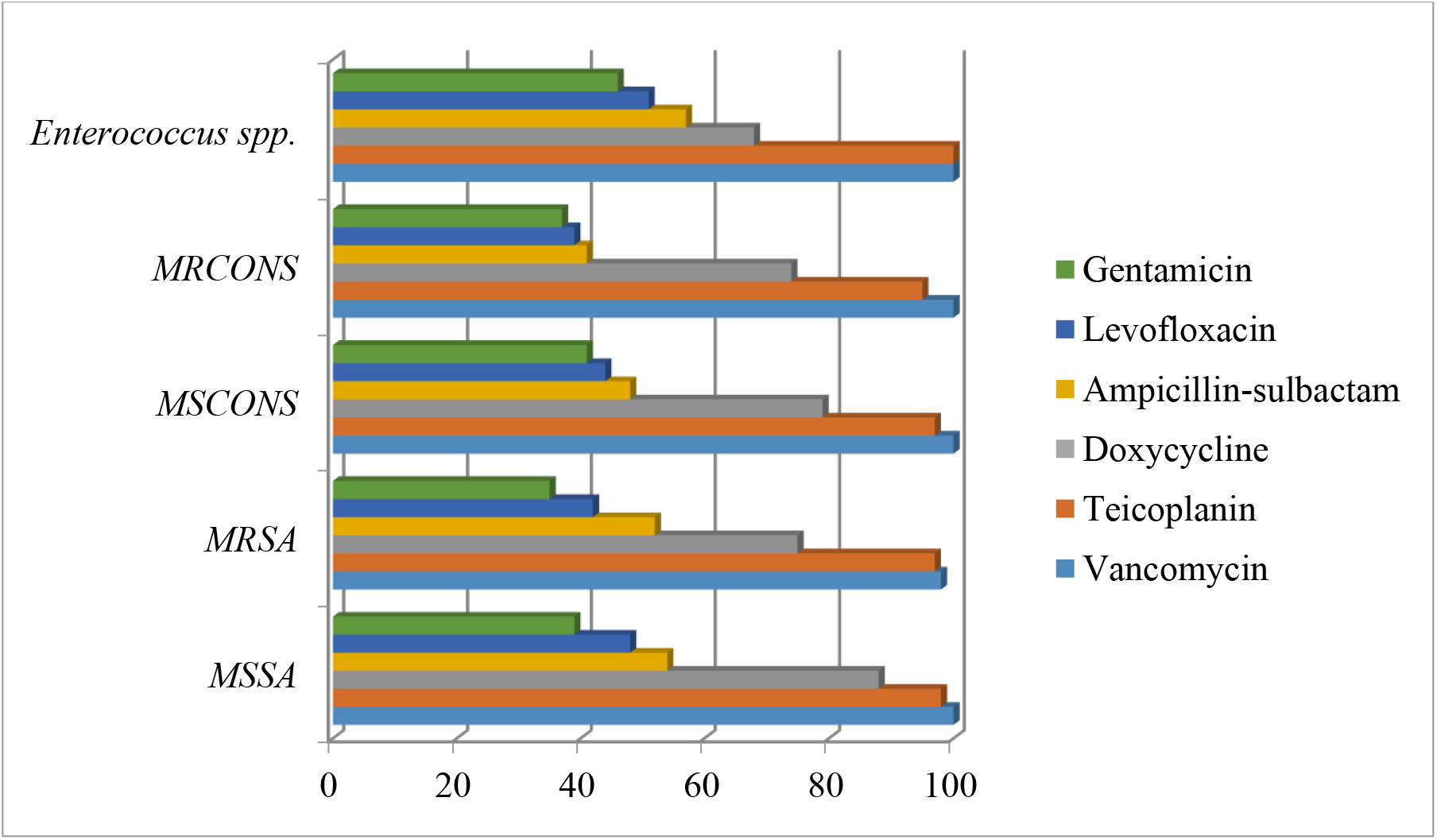
Antibiotic sensitivity pattern in Gram-positive isolates.

### Antibiotic usage

As per the WHO criteria of AWaRe class of antibiotics, maximum antibiotics were prescribed from the “watch” (48.02 %) and “reserve” categories (24.21 %). Only 20.7 % were given antibiotics from the “access” category while 7.07 % of the patients were not recommended any antibiotics as depicted in Fig 3 A & B.

**Fig 3:**
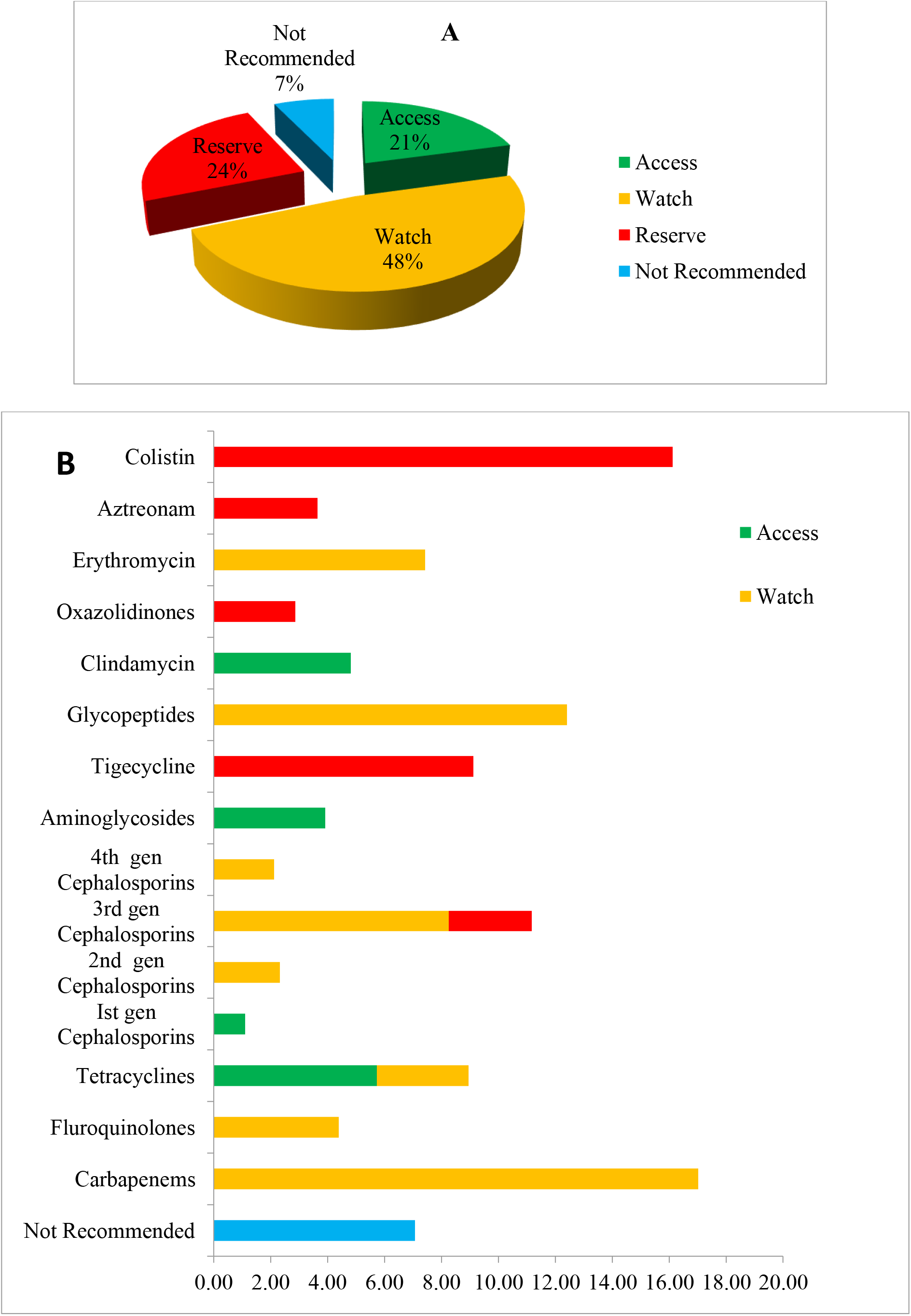
Illustration of antibiotics as per WHO Aware, Watch and Reserve (AWaRe) classification. (A) Total antibiotics usage in %. (B) Class of antibiotics usage in %.

### Clinical Outcomes

The two groups of COVID-19 positive patients with and without bacterial co-infections were compared in different clinical parameters and outcomes (Table 1). Patients with bacterial co-infections belonged to the older age group as compared to the patients without bacterial co-infections. No statistical significance was noted in terms of gender and other co-morbidities like hypertension, diabetes, CKD, CLD, COPD/asthma on comparing between the two groups. However; on comparing the clinical outcomes a significant difference was noted in terms of in-patient mortality, number of patients requiring intubation, use of ventilators and vasopressors. Patients with bacterial co-infections experienced a mortality rate of 39% as compared to non-co-infection group (17%). The requirement of ventilators was higher in patients with co-infection (35%) as compared to the non-co-infection group (21%). Similarly, a higher percentage of patients with bacterial co-infections (37%) needed intubation as compared to non-co-infection group (29%). Requirement of vasopressive drugs was more in patients with bacterial co-infections (32%) as compared to the patients with co-infection (14%).

**Table 1:**
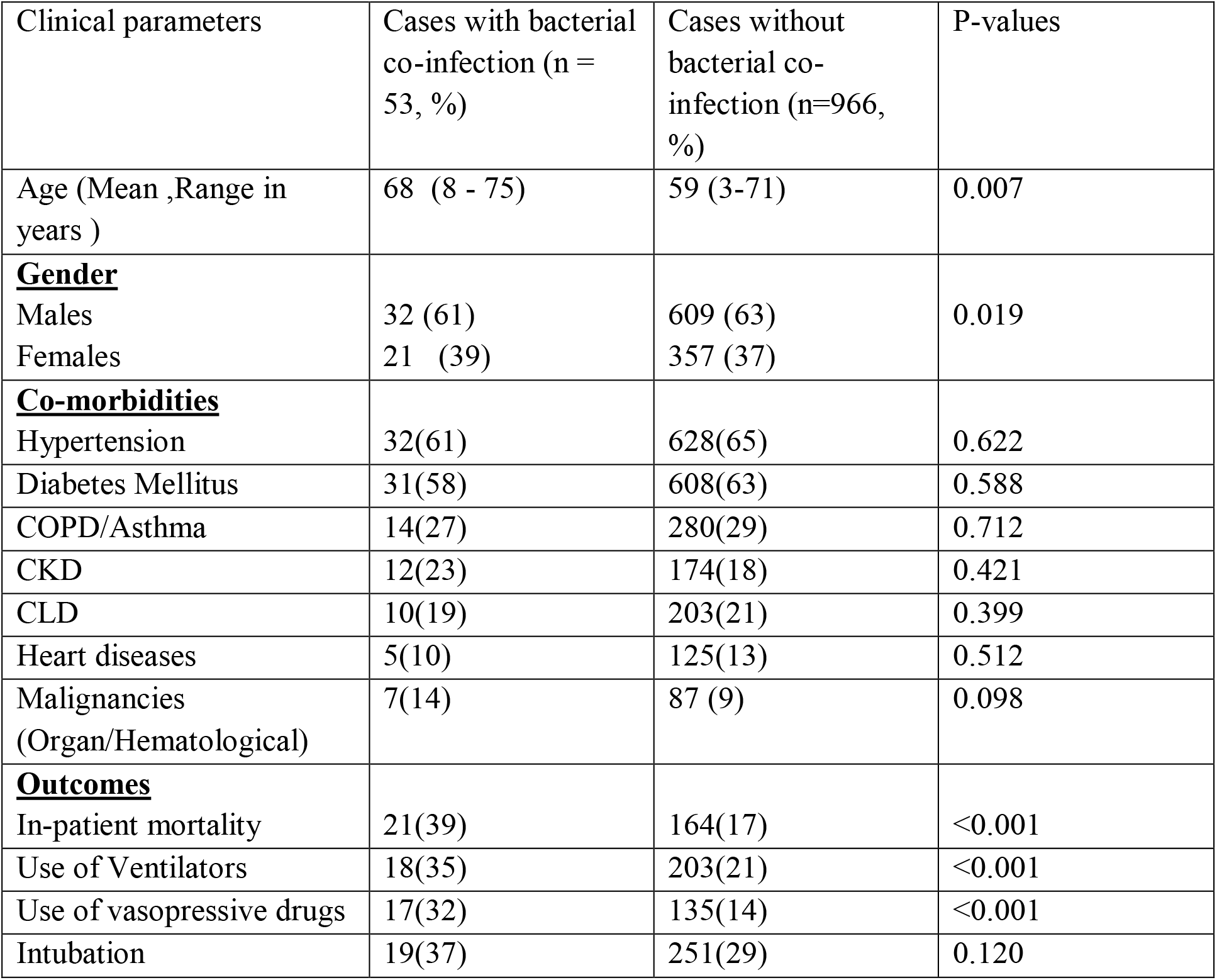
Comparison of co-morbidities and clinical outcomes in the two groups of COVID-19 patients (with and without bacterial co-infections).

| Clinical parameters | Cases with bacterial co-infection (n = 53, %) | Cases without bacterial co-infection (n=966, %) | P-values |
| --- | --- | --- | --- |
| Age (Mean ,Range in years ) | 68 (8 - 75) | 59 (3-71) | 0.007 |
| **<u>Gender</u>** |  |  |  |
| Males | 32 (61) | 609 (63) | 0.019 |
| Females | 21 (39) | 357 (37) |  |
| **<u>Co-morbidities</u>** |  |  |  |
| Hypertension | 32(61) | 628(65) | 0.622 |
| Diabetes Mellitus | 31(58) | 608(63) | 0.588 |
| COPD/Asthma | 14(27) | 280(29) | 0.712 |
| CKD | 12(23) | 174(18) | 0.421 |
| CLD | 10(19) | 203(21) | 0.399 |
| Heart diseases | 5(10) | 125(13) | 0.512 |
| Malignancies (Organ/Hematological) | 7(14) | 87 (9) | 0.098 |
| **<u>Outcomes</u>** |  |  |  |
| In-patient mortality | 21(39) | 164(17) | <0.001 |
| Use of Ventilators | 18(35) | 203(21) | <0.001 |
| Use of vasopressive drugs | 17(32) | 135(14) | <0.001 |
| Intubation | 19(37) | 251(29) | 0.120 |

## Discussion

SARS CoV-2 is a newly emerging virus which has led to a global pandemic in a span of only a few months. Its immunology, pathogenesis, clinical features and implications on the health care settings are yet to be fully understood. There is a lack of clinical research and data on the bacterial infections in these COVID-19 patients. Our study reported a bacterial co-infection rate of 5.2 % which is very well in agreement to other similar studies done in various parts of the world. A case series from Washington reported bacterial co-infection rate as 4.8%; while some studies from China also reported a rate of 5-9% [18–24]. A meta-analysis done by Lansbury et al., reported a bacterial co-infection rate of around 6.8% in hospitalized COVID-19 cases [25].

The current study stressed the prevalence of bacterial co-infections to be higher among the elderly age group (>65years). Several studies have emphasized the point of enhanced pathogenesis of COVID-19 in the elderly age group. A lot of factors like decreased mucociliary clearance, ciliary ultrastructural anomalies and immunosenescence play a key role in this. “Inflamm-aeging “or increased release of inflammatory mediators and cytokines leading to a cytokine surge is involved in tissue damage and multi-organ failure in such patients [26]. A higher proportion of co-morbidities like diabetes and hypertension were seen in the COVID cases in our study. This can be supported by the fact that diabetes mellitus itself down-regulates the immune system by decreasing the effective T-cell and neutrophils response [27]. It causes decreased phagocytosis, ineffective chemotaxis and decreased killing of invading microbes by neutrophils and macrophages leading to increased susceptibility to secondary bacterial infection [28]. Our study however failed to show any statistical significance for the association of various co-morbidities to the increase in bacterial co-infection rates among the COVID-19 cases. As such no role of increase in bacterial co-infection rates per se have been described so far in this sub-group of patients.

Extensive search of literature revealed that there is no available report of high rate of bacteremia and concurrent urinary tract infection (UTI) in COVID-19 patients. Ours is first such study revealing bacteremia (37%) and UTIs (31%) as the most common co-infections in COVID-19 patients. Other isolates were also isolated from respiratory (28%) and pus/aspirate (4%) samples in the current study. *E. coli, Klebsiella* and *Pseudomonas* were the predominant Gram-negative pathogens. One similar study in COVID-19 patients reported UTI in almost half of the patients (57%) [29]. It also reported *E. coli* and *Enterobacter cloacae* as the most commonly isolated pathogens. Another study by Lansbury et al., reported respiratory pathogens such as *Mycoplasma pneumoniae, P. aeruginosa*, and *Haemophilus influenza* as the most common isolates [25]. Increased prevalence of Gram-negative as well as Gram-positive pathogens in the current study can be attributed to the immune dysregulation leading to nosocomial infections and gut dysbiosis in COVID-19 positive patients. The inflammatory mediators disrupt the intestinal permeability leading to the leakage of gut microbes and associated metabolites into circulation. The leaked microbes and products via circulation migrate to organs including lungs and produce bacteremia, UTIs and various other infections [30]. Higher rates of diabetes in our group of patients also predispose to increase in secondary bacterial infections including UTIs [31]. Immune dysregulation in SARS-CoV-2 infection is characterized by lymphopenia, increased neutrophil–lymphocyte ratio, decreased NK-cells and CD8+T-cell activity, decreased regulatory T-cells and increased CD4+ to CD8+ ratio. Failure to eliminate virus due to inappropriate IFN response and decreased number and function of CD8+ and NK-cells further leads to virus induced tissue damage and makes the body more prone to secondary bacterial infections [31].

Colistin, Fosfomycin and Vancomycin proved to be some very effective drugs in treating bacterial infections in COVID-19 positive cases. Administration of antibiotics in this co-infection sub-group is vital to combat the ongoing bacterial infection in the form of blood stream, urinary and respiratory infections as well as to avoid the increased chances of acquiring secondary bacterial infections in such co-morbid patients. There was no significant past history of any antibiotic or antimicrobial usage in these groups of patients. Providing the correct and narrowed antibiotic coverage through proper antibiotic sensitivity testing will look after the ill effects caused by broader empiric treatment in such cases. As already highlighted in the study, most antibiotics prescribed were from the “watch” and “reserve” categories of WHO AWaRe classification. Antibiotic stewardship programmes will lead the path towards righteous treatment in COVID-19 infection group and will prevent the after effects of long term treatment.

Discussing the clinical outcomes of our study, we noticed a much higher rate of in-patient mortality in bacterial co-infection COVID-19 group (39%) as compared to the no infection group (17%). A similar study by Alvaro et al., reported a mortality rate of 50% in COVID-19 patients with concomitant bacterial infection [29]. While a study from Wuhan, China also reported similar mortality rates in this group of patients [20]. The increase in mortality rates may also be attributed to the older population involved with bacterial co-infections along with presence of other co-morbidities [32, 33]. The older population has weakened immune system due to immunosenescence and hence succumbs to the infections more easily than the younger ones. These people, therefore, more likely require interventions in the form of intubation, mechanical ventilation and use of vasopressive drugs to improve the outcome and increase their life expectancy. This fact was strongly suggested by our study where we noticed statistical significance in these COVID-19 cases with bacterial co-infection for the use of ventilator support, intubation and use of vasopressors as compared to the no infection group.

A lot of aspects still need to be explored in the current COVID-19 pandemic situation for the better understanding of its pathogenesis and control. Bacterial co-infections not necessarily but certainly may increase the mortality rates in COVID-19 positive patients. Antibiotic prescription as well as usage in this current situation must be properly guided through proper culture reports, sensitivity testing and stringent antibiotic stewardship programmes.

## Data Availability

The datasets used and/or analyzed during the current study are available from the corresponding author on reasonable request.

## Declarations

### Ethics approval

The study was approved by the institutional ethics committee of Sanjay Gandhi Post Graduate Institute of Medical Sciences under reference number 2020-100-EMP-EXP-16. Since, the study does not involve direct participation of human subjects so waiver of consent was granted.

### Consent to participate

Not applicable

### Author’s contributions

AP and CS conceptualized and supervised the study. AP, SS, SS and NF collected the samples, performed the testing and drafted the manuscript. AG, UG, SSP, AP and SS supervised the COVID-19 testing. AP and CS edited the manuscript.

